# LDLR Variant Classification for Improved Cardiovascular Risk Prediction in Familial Hypercholesterolemia

**DOI:** 10.1101/2023.12.28.23300621

**Authors:** Shirin Ibrahim, Merel L. Hartgers, Laurens F. Reeskamp, Linda Zuurbier, Joep Defesche, John J.P. Kastelein, Erik S.G. Stroes, G. Kees Hovingh, Roeland Huijgen

## Abstract

**Background:** Familial Hypercholesterolemia (FH) is a genetic disorder marked by high LDL cholesterol and an increased premature coronary artery disease (CAD) risk. Current dichotomous classification of LDL receptor gene (*LDLR*) variants may inadequately capture patient variability in LDL cholesterol levels and CAD risk. This study assessed a novel approach for assessing *LDLR* variant severity using variant-specific LDL cholesterol percentiles.

**Methods:** Participants of the Dutch FH cascade screening program were screened for 456 *LDLR* variants. Sex- and age-specific LDL cholesterol percentiles were computed for each *LDLR* variant carrier. These percentiles were used to calculate the mean LDL cholesterol percentile for each variant. Based on the variant-specific LDL cholesterol percentiles, carriers were grouped into the following strata: <75^th^, 75^th^-88th, 88^th^-92^nd^, 92^nd^-96.5^th^, 96.5^th^-98^th^, and ≥98^th^ percentile. Additionally, variants were categorized into class 1 (LDLR deficient) and non-class 1 (often LDLR defective) variants. CAD risk between carriers in the different strata and non-carriers was compared using a Cox proportional hazard model.

**Results:** Out of 35,257 participants, 12,485 (36%) *LDLR* variant carriers were identified. Carriers had a 5-fold higher CAD risk compared with non-carriers. Hazard ratios for CAD increased gradually from 2.2 (95%CI 0.97-5.0) to 12.0 (95%CI 5.5-24.8) across the strata. A 7.3-fold and 3.9-fold increased CAD risk was observed in carriers of class 1 and non-class 1 *LDLR* variants, respectively.

**Conclusions:** This study presents a refined approach for classifying *LDLR* variants based on their impact on LDL cholesterol levels, allowing for more precise, genotype-specific CAD risk estimation in FH patients compared with traditional methods.

## Introduction

Familial hypercholesterolemia (FH) is a common genetic disorder characterized by elevated levels of low-density lipoprotein (LDL) cholesterol caused by pathogenic variants in one of the three FH genes (LDL receptor gene [*LDLR*], apolipoprotein B gene [*APOB*], and proprotein convertase subtilisin kexin type 9 gene [*PCSK9*]), ultimately leading to premature morbidity and mortality due to atherosclerotic cardiovascular disease (ASCVD)^1–4^. To date, approximately 3000 FH-causing variants have been reported, with >90% of the monogenic FH cases being caused by pathogenic variants in *LDLR*^5, 6^. The effect of the different FH-causing variants on residual LDLR activity varies widely, leading to a broad spectrum of LDL cholesterol levels and subsequent risk of coronary artery disease (CAD) among FH patients^6–8^. This diversity in LDLR functionality is often simply classified into two categories – receptor “deficient” (residual LDLR activity <2%) or receptor “defective” (residual LDLR activity 2-25%) – thereby ignoring the wide range of impact these variants can have^9,10^. However, understanding the broad spectrum of LDL cholesterol levels and CAD risk stemming from the diverse effects of FH-causing variants is crucial, as it directly influences patient outcomes.

Pathogenicity evaluation of newly identified variants in one of the FH genes typically relies on *in silico* or *in vitro* analyses. However, *in vitro* studies are laborious and infrequently performed, while *in silico* predictions often do not consistently match clinical observations^7, 11^. As a consequence, the actual pathogenicity of many variants is unknown. A more clinically relevant classification of FH-causing variants might be achieved by considering LDL cholesterol levels and CAD risk caused by the specific variants *in vivo*, rather than the traditional method focused on receptor deficiency or defectiveness.

The aim of the current study was to assess the severity of pathogenic variants in *LDLR* based on LDL cholesterol levels in a large cohort of heterozygous FH patients. Furthermore, the correlation between variant-specific LDL cholesterol levels (as indicators of LDLR functionality or loss thereof) and CAD risk was examined, aiming to enhance the accuracy of cardiovascular risk prediction in FH patients.

## Methods

### Study population

The present study included carriers of an FH-causing variant who participated in the Dutch national FH cascade screening program^12, 13^. Between January 1994 and April 2013, this FH screening initiative resulted in genetic testing in over 60,000 individuals, identifying more than 28,000 carriers of FH-causing variants via a cascade screening method. Index patients were excluded from the current study to avoid clinical sampling bias. In addition, homozygous FH patients and individuals carrying pathogenic variants in *APOB* or *PCSK9* were also excluded. This study was approved by the Institutional Review Board of Amsterdam UMC, the Netherlands, and complied with the Declaration of Helsinki. All participants gave written informed consent.

### Study procedures

Prior to 2003, lipid data were not consistently available for all participants of the program. From 2003 onwards, the screening program used the LDX-analyzer to measure lipid levels. LDL cholesterol levels were estimated by the Friedewald equation, unless triglycerides levels were above 400 mg/dl. Genetic testing was performed through Sanger sequencing or Multi Ligand Probe Amplification (MLPA) of a specific FH-causing variant or copy number variation (CNV) in *LDLR*.

### Variant classification

For each participant, a sex- and age-specific LDL cholesterol percentile was calculated, using previously established LDL cholesterol percentiles in healthy controls, who were not receiving lipid-lowering therapy (LLT)^8^. In participants who only had on-treatment LDL cholesterol levels available, pre-treatment levels were calculated based on the average LDL cholesterol reduction achieved in trials with a given (combination) therapy^14–17^. Subsequently, for each specific variant in *LDLR*, the mean LDL cholesterol percentile was calculated based on LDL cholesterol percentiles of all carriers of that variant. Next, strata were formed according to the variant-specific LDL cholesterol percentiles; <75^th^, 75^th^-88^th^, 88^th^-92^nd^, 92^nd^-96.5^th^, 96.5^th^-98^th^ and ≥98th percentile. The chosen cutoffs were strategically selected. In previous research, the 75^th^ percentile served as a benchmark to distinguish individuals or LDLR variants with a modest or negligible FH phenotype when below this threshold. Variants with numerous carriers below this threshold were scrutinized for potential non-pathogenicity^7, 11, 18^. The other thresholds were chosen to achieve groups with a sufficient amount of carriers

In addition, variants were also categorized using the widely used methodology based on LDLR functionality^19^. According to this methodology, variants can be classified as class 1 variants and non-class 1 variants. Class 1 variants are generally the most severe variants because no LDLR protein can be synthesized at all from the mutated allele, representing a complete loss of function of the LDLR defined by an LDLR activity <2% (receptor deficient or null variants). In non-class 1 variants there is often some residual LDLR activity (LDLR defective variants), denoting partial function with LDLR activity of 2-25%. Thus, class 1 variants are generally considered more severe due to absent of LDLR protein synthesis and thus absent of residual LDLR activity. In contrast, non-class 1 variants are mostly receptor defective and are in general considered less severe.

### Risk stratification of coronary artery disease in LDLR variant carriers

All study participants were alive at time of genetic testing and received no follow-up after participating in the cascade screening program. CAD was defined as a history of prior non-fatal cardiac events (myocardial infarction, percutaneous coronary intervention, or coronary artery bypass graft) at the time of testing. The duration of CAD event-free survival was calculated from birth until the first occurrence of a CAD event or until January 1^st^, 1990, whichever came first. This cutoff date was selected because statins became available in the Netherlands in 1990, and data on CAD events after this year could be influenced by statin use.

### Statistical analysis

Differences in normally distributed variables were compared using unpaired Student t-tests. Dichotomous variables were analyzed using chi-square tests. A Cox proportional hazards model adjusted for gender, age, hypertension, diabetes, smoking habits, and body mass index, was applied to evaluate the risk of CAD in different *LDLR* variant carriers, with unaffected relatives serving as the reference group. A two-sided p-value <0.05 was considered significant. Data analysis was performed using SPSS for Windows version 23 (Chicago, IL).

## Results

### Study population

A total of 40,447 individuals underwent FH cascade testing between 1994 and 2010. After exclusion of carriers of an FH-causing variant in *APOB* and *PCSK9* (n = 5190), and *LDLR* variant carriers without any available LDL cholesterol measurements for that specific variant (n = 190), a study population of 35,067 subjects remained, who were screened for the presence of the specific *LDLR* variant identified in the index patient of their family (**Figure 1**). The mean year of screening was 2005. A total of 456 distinct *LDLR* variants were considered pathogenic at the time genetic cascade screening was performed (**Supplementary Table S1)**. Of the 35,067 subjects, 12,485 (36%) were heterozygous carriers of an FH-causing variant in *LDLR* and 22,582 (64%) were non-carriers (unaffected family members). Clinical characteristics of these two groups are shown in **Table 1**. Compared to the non-carriers, FH patients had a lower mean age (38.0±20.0 vs. 43.7±19.0 years), a lower Body Mass Index (24.0±4.9 vs. 25.0±4.6 kg/m^2^), and a lower prevalence of smoking, diabetes and hypertension (28% vs. 34%, 2.2% vs. 3.2% and 8.5% vs. 12%, respectively). Moreover, FH patients had a higher mean LDL cholesterol than non-carriers (162.4±54.1 vs. 112.1±36.7 mg/dl) and a higher prevalence of CAD (6.9% vs. 3.6%).

**Figure 1.**
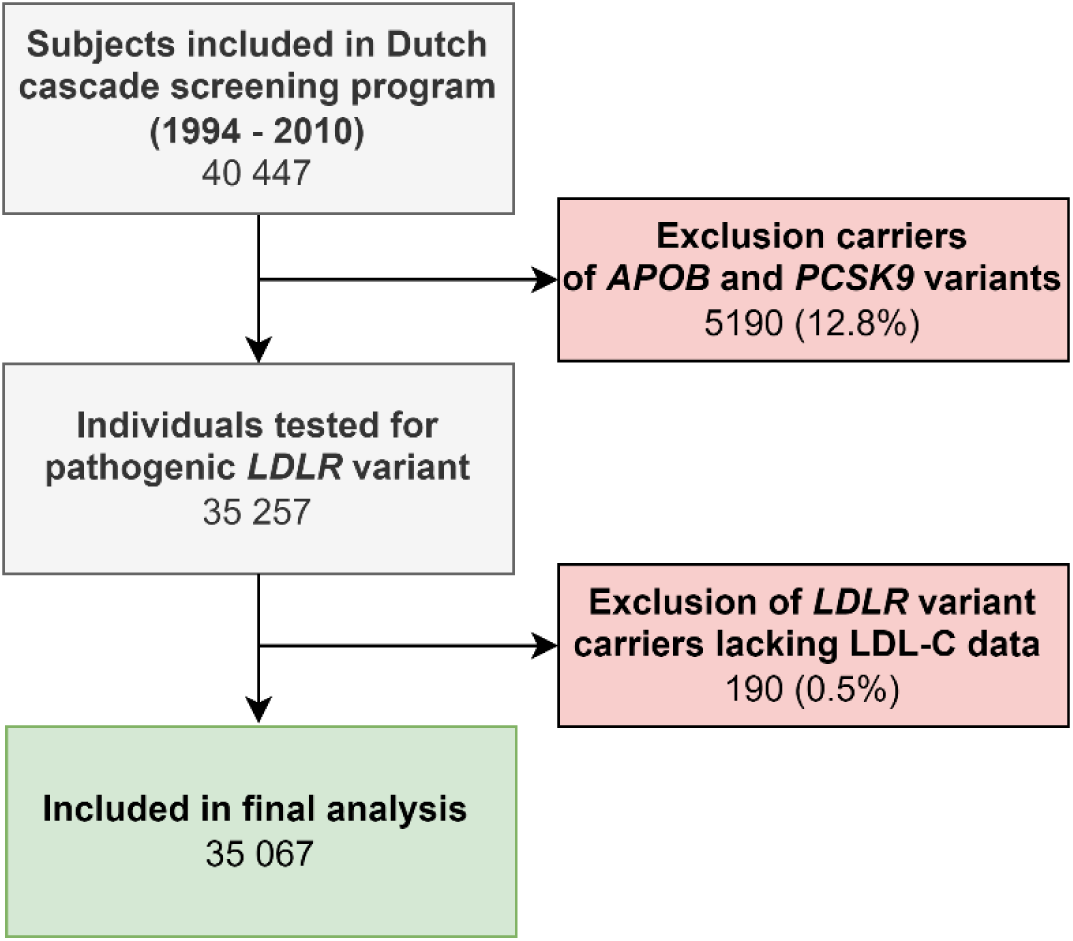
Flowchart of participant selection. The study population originated form the Dutch familial hypercholesterolemia cascade screening program. *APOB*, apolipoprotein B gene; LDL-C, low-density lipoprotein cholesterol; LDLR, low-density lipoprotein receptor; *PCSK9*, proprotein convertase subtilisin kexin type 9 gene.

**Table 1.**
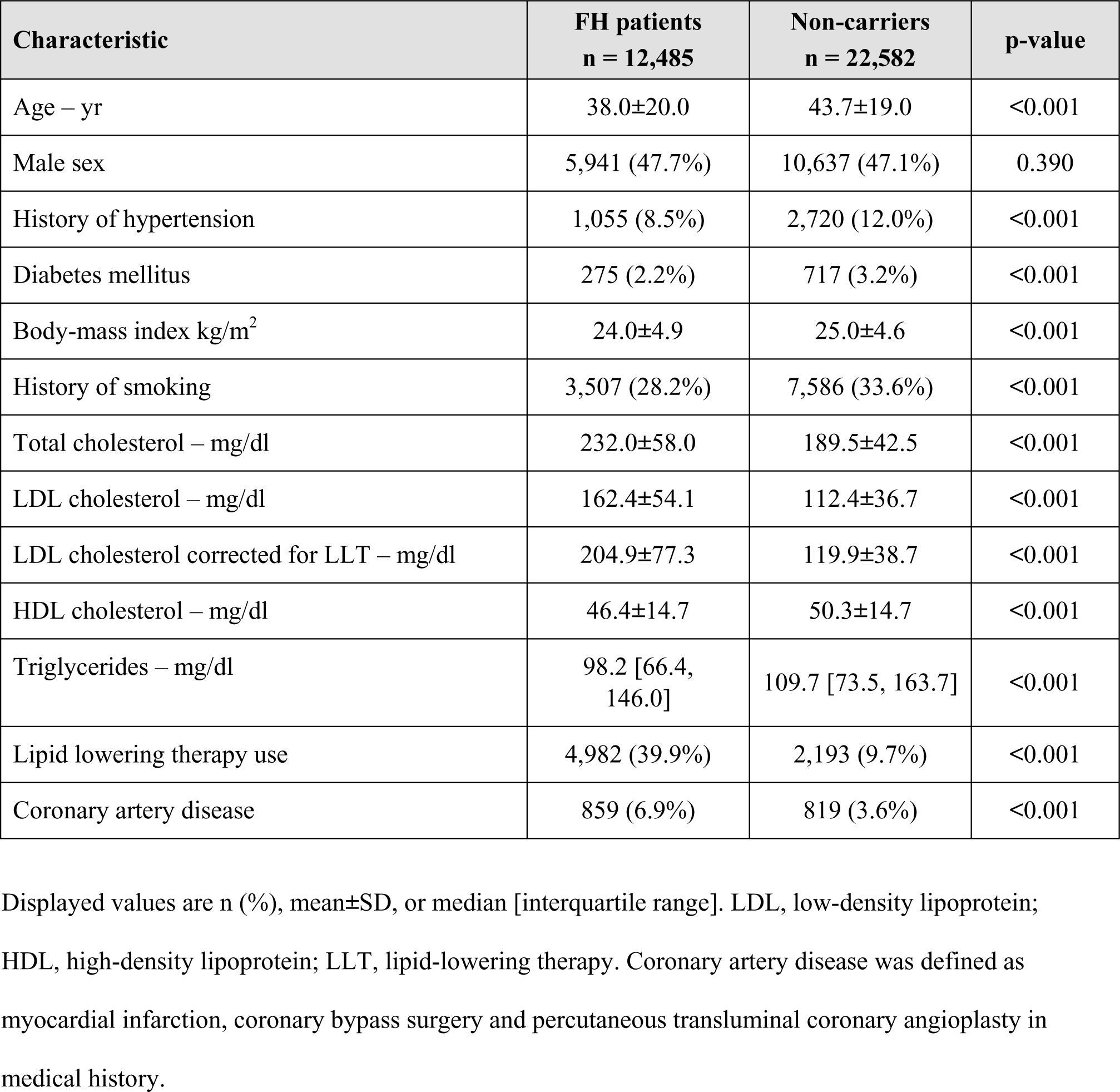
Participant characteristics.

| Characteristic | FH patients<br>n = 12,485 | Non-carriers<br>n = 22,582 | p-value |
| --- | --- | --- | --- |
| Age – yr | 38.0±20.0 | 43.7±19.0 | <0.001 |
| Male sex | 5,941 (47.7%) | 10,637 (47.1%) | 0.390 |
| History of hypertension | 1,055 (8.5%) | 2,720 (12.0%) | <0.001 |
| Diabetes mellitus | 275 (2.2%) | 717 (3.2%) | <0.001 |
| Body-mass index kg/m<sup>2</sup> | 24.0±4.9 | 25.0±4.6 | <0.001 |
| History of smoking | 3,507 (28.2%) | 7,586 (33.6%) | <0.001 |
| Total cholesterol – mg/dl | 232.0±58.0 | 189.5±42.5 | <0.001 |
| LDL cholesterol – mg/dl | 162.4±54.1 | 112.4±36.7 | <0.001 |
| LDL cholesterol corrected for LLT – mg/dl | 204.9±77.3 | 119.9±38.7 | <0.001 |
| HDL cholesterol – mg/dl | 46.4±14.7 | 50.3±14.7 | <0.001 |
| Triglycerides – mg/dl | 98.2 [66.4,<br>146.0] | 109.7 [73.5, 163.7] | <0.001 |
| Lipid lowering therapy use | 4,982 (39.9%) | 2,193 (9.7%) | <0.001 |
| Coronary artery disease | 859 (6.9%) | 819 (3.6%) | <0.001 |

Displayed values are n (%), mean±SD, or median [interquartile range]. LDL, low-density lipoprotein; HDL, high-density lipoprotein; LLT, lipid-lowering therapy. Coronary artery disease was defined as myocardial infarction, coronary bypass surgery and percutaneous transluminal coronary angioplasty in medical history.

### Variant-specific LDL cholesterol percentile strata

Clinical characteristics of the groups stratified according to the variant-specific LDL cholesterol percentiles are shown in **Table 2**. Subjects in the lowest LDL cholesterol percentile stratum were older compared to those in the highest stratum (40.4±20.2 years vs. 37.7±19.4 years), and were less often treated with LLT at the time of genetic diagnosis (16.1% vs. 47.7%, OR 5.3 [95%CI 4.3 – 6.5], p<0.0001). Carriers of class 1 variants were identified across all LDL cholesterol strata; 7.9% of the patients in the lowest LDL cholesterol stratum and 53% of the patients in the highest LDL cholesterol stratum carried a class 1 variant (**Table 2**).

**Table 2.** Characteristics of carriers in the variant-specific LDL cholesterol percentile strata.

| Characteristic | <75<sup>th</sup><br>percentile<br>n = 983 | 75<sup>th</sup>-88<sup>th</sup><br>percentile<br>n = 972 | 88<sup>th</sup>-92<sup>nd</sup><br>percentile<br>n = 3,245 | 92<sup>nd</sup>-96.5<sup>th</sup><br>percentile<br>n = 2,793 | 96.5<sup>th</sup>-98<sup>th</sup><br>percentile<br>n = 3,377 | >98<sup>th</sup><br>percentile<br>n = 1,115 |
| --- | --- | --- | --- | --- | --- | --- |
| Age – yr | 40.0±20.0 | 40.0±24 | 38±19 | 37±19 | 36±20 | 36±19 |
| Male sex | 445 (45.3) | 457 (47.0) | 1,560 (48.1) | 1,381 (49.4) | 1,594 (47.2) | 504 (45.2) |
| History of hypertension | 114 (11.6) | 119 (12.2) | 231 (7.1) | 247 (8.8) | 259 (7.7) | 85 (7.6) |
| Diabetes mellitus | 43 (4.3) | 43 (4.4) | 45 (1.4) | 58 (2.1) | 60 (1.8) | 26 (2.3) |
| Body-mass index kg/m<sup>2</sup> | 24±5.0 | 24±4.8 | 24±4.5 | 24±4.8 | 23±5.0 | 24±5.2 |
| History of smoking | 320 (32.6) | 297 (30.6) | 905 (27.9) | 783 (28.0) | 877 (26.0) | 325 (29.1) |
| Total cholesterol – mg/dl | 193.3±46.4 | 216.6±50.3 | 228.2±50.3 | 232.0±58.0 | 247.5±61.9 | 251.4±61.9 |
| LDL cholesterol – mg/dl | 119.9±38.7 | 143.1±46.4 | 154.7±46.4 | 166.3±54.1 | 177.9±58.0 | 181.7±58.0 |
| LDL cholesterol corrected for LLT – mg/dl | 127.6±42.5 | 166.3±58.0 | 193.3±61.9 | 216.6±73.5 | 239.8±81.2 | 251.4±73.5 |
| HDL cholesterol – mg/dl | 46.4±13.9 | 46.4±14.7 | 46.4±15.1 | 46.4±13.9 | 46.4±13.5 | 46.4±15.9 |
| Triglycerides – mg/dl | 113.3 [74.3,<br>166.4] | 100.9 [68.1,<br>151.3] | 95.6 [66.4,<br>145.1] | 93.8 [62.8,<br>138.9] | 97.4 [65.5,<br>143.4] | 100.9 [67.3,<br>142.5] |
| Lipid lowering therapy use | 158 (16.1) | 277 (28.5) | 1,064 (32.8) | 1,317 (47.2) | 1,634 (48.4) | 532 (47.7) |
| Carriers of class 1 variants | 78 (7.9) | 58 (6.0) | 70 (2.2) | 1,231 (44.1) | 2,152 (63.7) | 592 (53.1) |

Displayed values are n (%), mean±SD, or median [interquartile range]. LDL, low-density lipoprotein; HDL, high-density lipoprotein; LLT, lipid-lowering therapy.

### Risk stratification of coronary artery disease in LDLR variant carriers

To determine the event-free survival in the era before statins were widely used, data from the 31,217 subjects born before 1990 were used exclusively. Carriers of an FH-causing variant had a 5-fold higher CAD risk compared with non-carriers (HR 5.0 [95%CI 4.1 to 6.0]). CAD risk increased across the different variant-specific LDL cholesterol percentile strata, with LDL cholesterol percentiles of <75^th^, 75^th^-88^th^, 88^th^-92^nd^, 92^nd^-96.5^th^, 96.5^th^-98^th^, and >98^th^ being associated with HR’s of 2.2 (95%CI 0.97 to 5.0), 2.5 (95%CI 1.3 to 5.0), 3.8 (95%CI 2.6 to 5.7), 4.1 (95%CI 2.9 to 5.9), 8.8 (95%CI 6.2 to 13), and 12 (95%CI 5.5 to 24.8), respectively (**Figure 2**). In comparison, categorizing the variants based on LDLR functionality, non-class 1 *LDLR* variant carriers had a 3.9-fold (95%CI 3.0 to 4.9) higher CAD risk than their unaffected relatives and carriers of a class 1 *LDLR* variant had a 7.3-fold (HR, 95% CI: 5.4-9.3) higher CAD risk compared with their unaffected relatives.

**Figure 2.**
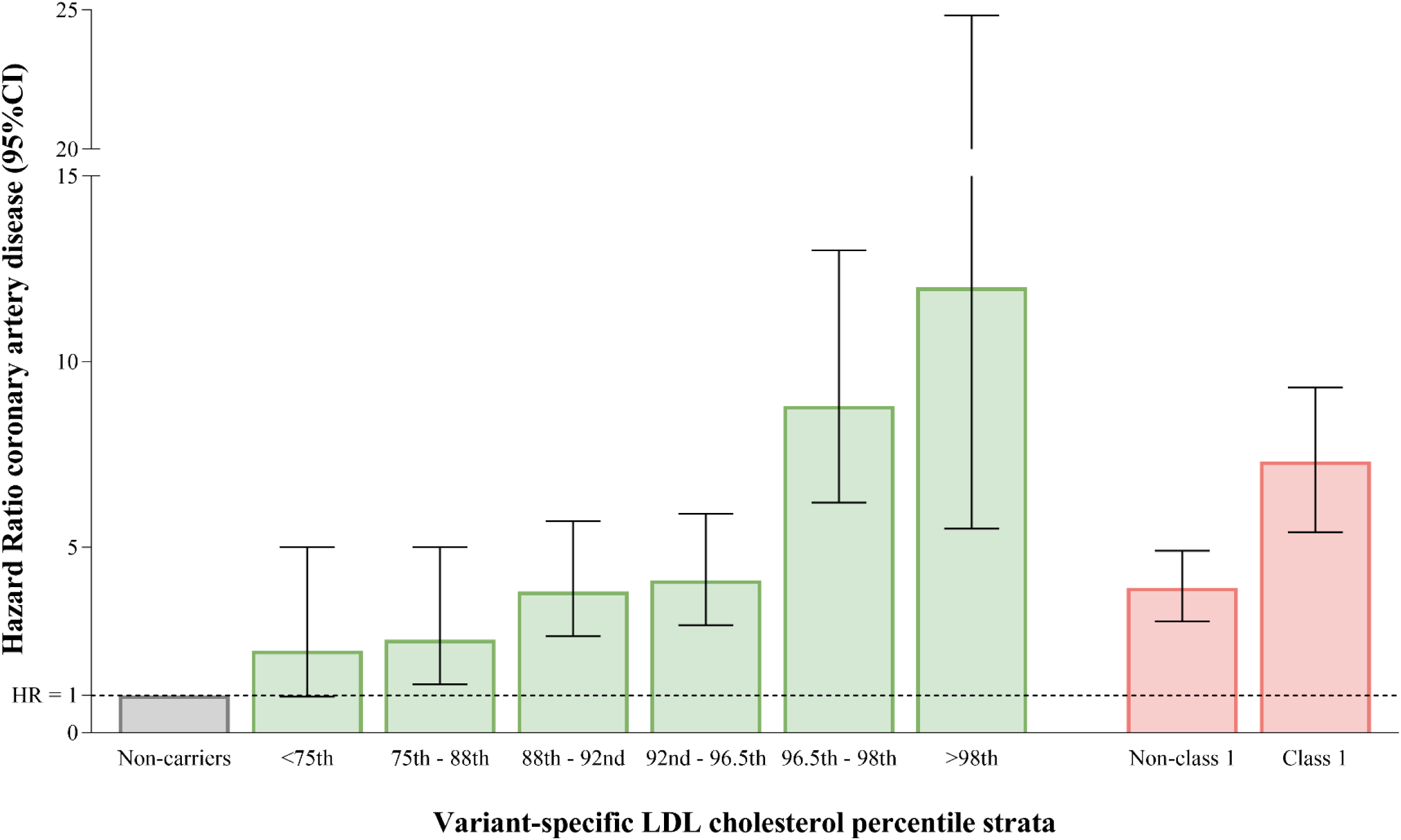
Coronary artery disease across the variant-specific LDL cholesterol percentile strata in pre-statin era. A Cox proportional hazard model, adjusted for age, sex, hypertension, diabetes, body mass index and history of smoking, was used to compare coronary artery disease (CAD) risk between carriers and non-carriers. The non-carriers served as a reference group (hazard ratio = 1). CAD was defined as a history of myocardial infarction, coronary bypass surgery and percutaneous transluminal coronary angioplasty. The analysis period, defined as the pre-statin era, spaned from birth to the first CAD event or until January 1, 1990, whichever occured first. The left side of the figure presents *LDLR* variant classification based on the mean percentile of LDL cholesterol for each variant (green). The right side presents *LDLR* variant classification into class 1 (receptor deficient) and non-class 1 (typically receptor defective) variants (red). LDL, low-density lipoprotein.

## Discussion

In this study, a novel method for classifying *LDLR* variants based on their impact on sex- and age-specific LDL cholesterol levels reveals a refined spectrum of CAD risk among FH patients, ranging from 2.2-fold to 12-fold compared to non-carriers. The conventional dichotomous classification of class 1 and non-class 1 variants was associated with HR’s of 3.9 and 7.3, respectively. The more refined risk stratification based on variant-specific LDL cholesterol percentiles offers clinicians a valuable tool for personalized risk assessment, potentially enabling more precise management strategies for FH patients.

In this large-scale study involving over 35,000 individuals screened for pathogenic *LDLR* variants, it was observed that carriers of an FH-causing variant face a 5-fold increased CAD risk compared to unaffected relatives. This risk varies significantly depending on the specific *LDLR* variant, with FH patients that carry variants associated with extreme LDL cholesterol levels (>98^th^ percentile for sex and age) demonstrating a 5-fold higher risk compared to carriers of variants leading to LDL cholesterol levels <75th percentile. Notably, mean off-treatment LDL cholesterol levels ranged from 98.2 mg/dl for carriers of the most modest *LDLR* variants to 355.0 mg/dl for carriers of the most severe variants. This variability in LDL cholesterol levels has substantial implications for CAD risk, emphasizing the importance of considering the total exposure to a genetically defined LDL cholesterol burden over time.

The insights from this study align with earlier large cohort studies employing exome sequencing for rare *LDLR* variants^20–22^. For example, Khera et al. reported similar findings in their study involving 26,050 individuals, where a 4-fold increased CAD risk was observed among carriers of an FH-causing variant in general and a 10-fold increase for those with class 1 *LDLR* variants^20^. The present study’s LDL cholesterol percentile-based approach identified severe *LDLR* variants posing a high CAD risk, which paralleled the risk levels observed in Khera et al.’s study for strictly defined class 1 variants. For the majority of variants, the residual LDLR function remains, however, uncertain. These variants without established LDLR function exhibit a considerable variation in FH phenotype. The methodology used in this study, using LDL cholesterol levels (corrected for sex and age) as an indicator of LDLR functionality, provides a useful tool for estimating residual receptor activity, which is especially useful in categorizing defective *LDLR* variants into specific risk categories. This is a notable improvement compared with *in vitro* studies and *in silico* tools that have limited applicability and accuracy^7, 11^.

The present study introduces a novel method for classifying the severity of pathogenic variants that enhances the precision of CAD risk stratification in FH. The findings highlight that with an LDL percentile-based classification, clinicians may be better suited to personalize the risk estimates of individual patients, unlike the conventional classifications based solely on receptor-deficient or receptor-defective models. This approach may help provide accurate patient counseling, initiate timely intervention in high-risk individuals, and decide on genetic cascade screening within families, especially when dealing with *LDLR* variants with a great impact on LDL cholesterol percentile.

### Limitations

In interpreting the findings of this study, several limitations must be considered. First, the risk estimates are based on a longitudinal analysis. Nonetheless, this study included an unprecedented large cohort of FH-variant carriers which enabled precise estimation of effects, especially since LDL cholesterol levels in non-carriers of the same families were used as the reference. Second, the analysis focused on non-fatal CAD events that occurred before the introduction of statins in 1990. This approach was chosen to minimize confounding from statin use, which is commonly prescribed to FH patients regardless of their CAD history. However, by focusing only on pre-1990 non-fatal events, there is a potential underestimation of the impact of the genetic variants. Moreover, the study included patients who had survived a non-fatal CAD event for several years, sometimes up to 15 years or more, leading to a potential survival bias that might have affected carriers and non-carriers differently. To address this selective survival bias, relative CAD risk instead of absolute CAD risk was reported. Lastly, this study’s variant classification method based on LDL cholesterol percentiles, may not be universally applicable. In many regions, where FH is primarily clinically diagnosed based on physical signs, family history, and lipid levels, and where genetic cascade screening is not standard practice, identifying a reference population of unaffected relatives is challenging, which makes the determination and calculation of LDL cholesterol percentiles for variant carriers complex.

## Conclusions

In conclusion, the approach presented in this study for classifying *LDLR* variants based on their impact on LDL cholesterol levels allows for a more precise genotype-specific CAD risk estimation in FH patients compared with traditional methods. This refined tool for CAD risk assessment marks a significant step towards more tailored and effective care in FH.

## Acknowledgements

We would like to express our sincere gratitude to participants of the genetic cascade-screening program for FH.

## Conflict of interest

**LFR** is cofounder of Lipid Tools and reports speakers fee from Ultragenyx, Novartis, and Daiichi Sankyo. **JJPK** is part time CSO of New Amsterdam Pharma and part time CMO of Staten Biotech and a consultant to AgBio, Scribe, Cincor, CSL Behring, CiVi Biopharma, Draupnir, Esperion Therapeutics, Madrigal, Menarini, Omeicos, Silence Therapeutics, Sirnaomics, TTxD and North Sea Therapeutics. **GKH** reports research grants from the Netherlands Organization for Scientific Research (vidi 016.156.445), CardioVascular Research Initiative, European Union and the Klinkerpad fonds, institutional research support from Aegerion, Amgen, AstraZeneca, Eli Lilly, Genzyme, Ionis, Kowa, Pfizer, Regeneron, Roche, Sanofi, and The Medicines Company; speaker’s bureau and consulting fees from Amgen, Aegerion,Sanofi, and Regeneron until April 2019 (fees paid to the academic institution); and part-time employment and stock holder of at Novo Nordisk A/S, Denmark since April 2019. **ESGS** has received ad-board/lecturing fees, paid to the institution, from: Amgen, Sanofi, Astra-Zeneca, Esperion, Daiichi Sankyo, NovoNordisk, Ionis/Akcea, Amarin. The other authors report no disclosures.

## Data availability statement

The data used in this study are available upon reasonable request from the corresponding author.

## Notes

### Clinical Trial

Not applicable.

### Funding Statement

No external funding was received.

### Author Declarations

This study was approved by the Institutional Review Board of Amsterdam UMC, the Netherlands, and complied with the Declaration of Helsinki. All participants gave written informed consent.

